# Evaluating the Accuracy of Longitudinal Strain from Poor-Quality Echocardiograms in Identifying STEMI Culprit Vessels

**DOI:** 10.1101/2025.03.03.25323187

**Authors:** Lark Steafo, Athanasios Rempakos, Julie George, Joel Skaistis

## Abstract

**Purpose:** Longitudinal strain is a valuable echocardiographic marker, yet its reliability in poor-quality images remains uncertain. Poor image quality is common in ST-segment elevation myocardial infarction (STEMI) patients, raising concerns about the utility of strain analysis in such cases. This study evaluated the accuracy of longitudinal strain in predicting the culprit vessel in STEMI patients with suboptimal echocardiographic imaging.

**Methods:** A retrospective analysis of 50 consecutive STEMI patients was performed using echocardiograms acquired during index hospitalizations. Studies with discernable basal insertion points and apical caps were included, despite limited visualization of other segments. Longitudinal strain was quantified using the 4AS parameter, derived with TomTec LV AutoStrain software, and compared against clinical interpretations performed without strain analysis. Culprit vessel identification by strain and clinical interpretation was validated against coronary angiography findings.

**Results:** The 4AS parameter correctly identified the culprit vessel in 48 of 50 cases (96%), significantly outperforming routine clinical interpretation, which was correct in 35 cases (70%; p = 0.0022). Strain performance was robust across varying image quality levels and coronary territories.

**Conclusions:** Longitudinal strain, as measured by 4AS, accurately identified STEMI culprit vessels despite poor echocardiographic image quality, surpassing standard clinical interpretations. These findings suggest that strain analysis retains diagnostic utility even with low-quality images, supporting its integration into routine workflows for acute cardiac care.

---

Acute myocardial infarction is a major contributor to morbidity and mortality, necessitating early and accurate assessment of myocardial injury for risk stratification and management. Traditional echocardiographic markers, such as left ventricular ejection fraction (LVEF), have been used to assess cardiac function. However, emerging studies have suggested that global longitudinal strain (GLS) provides further insight into myocardial injury. Studies have demonstrated that GLS is significantly linked with infarct size in the early stages of acute myocardial infarction, especially in patients with preserved LVEF [1]. Furthermore, GLS has been shown to outperform LVEF in assessing infarct size following percutaneous coronary intervention in patients with ST elevation myocardial infarction [2]. Beyond estimating infarct size, GLS has also been shown to serve as a valuable prognostic marker, predicting major cardiac adverse events, including cardiac death, re-infarction, and hospitalization for heart failure, comparable to LVEF in the acute phase and exceeding LVEF predictive capability after 10 days [3]. However, it remains unclear whether longitudinal strain is only of value when performed on high quality echocardiographic images with well defined endocardial borders or whether it still conveys valuable information when performed in cases of poor image quality. Strain is typically measured using artificial intelligence (AI) tools that automatically detect the myocardium from apical views. Conventional wisdom suggests that AI derived strain analysis might be unreliable when performed on poor quality images, following the “garbage in, garbage out” adage. This raises concerns about the applicability of GLS in a real world setting, where suboptimal image quality is common, especially in STEMI patients, due to their clinical instability, discomfort, and restrictions in positioning following catheterization procedures. This study aimed to evaluate the accuracy of longitudinal strain in identifying the culprit vessel in a cohort of patients presenting with ST-segment elevation myocardial infarction (STEMI) who had poor quality echocardiographic images. By investigating whether GLS remains a valuable diagnostic marker under these conditions, this study aims to highlight the robustness of this tool in clinical practice to improve patient outcomes, especially when imaging limitations exist.

The echocardiograms of 50 consecutive STEMI patients at a large tertiary hospital were retrospectively analyzed. Echocardiograms were acquired during the STEMI index hospitalizations with the majority performed after reperfusion. Echocardiograms were reviewed by a Level 3 echocardiographer (JS), and studies were included if the basal insertion points and apex were discernible throughout the cardiac cycle in all apical views. Image quality was graded on a three-point scale as somewhat limited, intermediate and very limited, corresponding to studies with less than 25% of segments not visualized, 25-50% of segments not visualized and >50% of segments not visualized, respectively, as seen in Figure 1.

**Fig 1.**
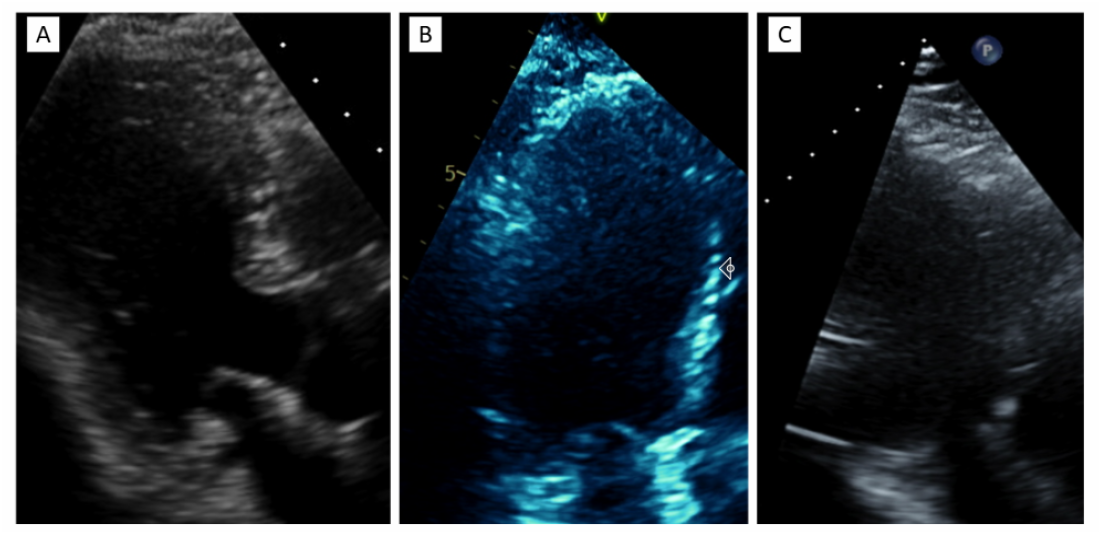
Examples of “somewhat limited” (A), “intermediate” (B), and “very limited quality” (C) 3 chamber views

TomTec LV AutoStrain (TomTec Imaging Systems GmbH, Munich, Germany, version TTA 2.41) was used to quantify global longitudinal strain. The culprit vessel was identified according to the myocardial segment with the lowest 4AS score [4]. The 4AS score averages the longitudinal strain values of the 4 contiguous segments with the lowest absolute strain values. Absolute values less than 15% were considered abnormal. No more than two mid chamber segments may be incorporated into the score. Myocardial segments from the 17 segment AHA model were assigned to coronary territories according to figure 2. The lateral wall was assigned upon review of the coronary angiogram. In cases with large RCA PLV branches, the inferolateral wall was assigned to the RCA and in cases with large diagonal branches, the anterolateral wall was assigned to the LAD. Strain interpretation was performed while blinded to the coronary angiogram. 4AS predictions were compared against routine clinical interpretation, which incorporated contrast imaging but not strain analysis and was performed at the time of image acquisition by the treatment team. The culprit vessel for each case was confirmed through review of coronary angiograms with the culprit vessel defined as the vessel with the lowest pre-revascularization TIMI flow grade. Chi-square tests were used to compare correct identification rates between LV AutoStrain and clinical interpretation with Fisher’s exact tests for small sample sizes.

**Fig 2.**
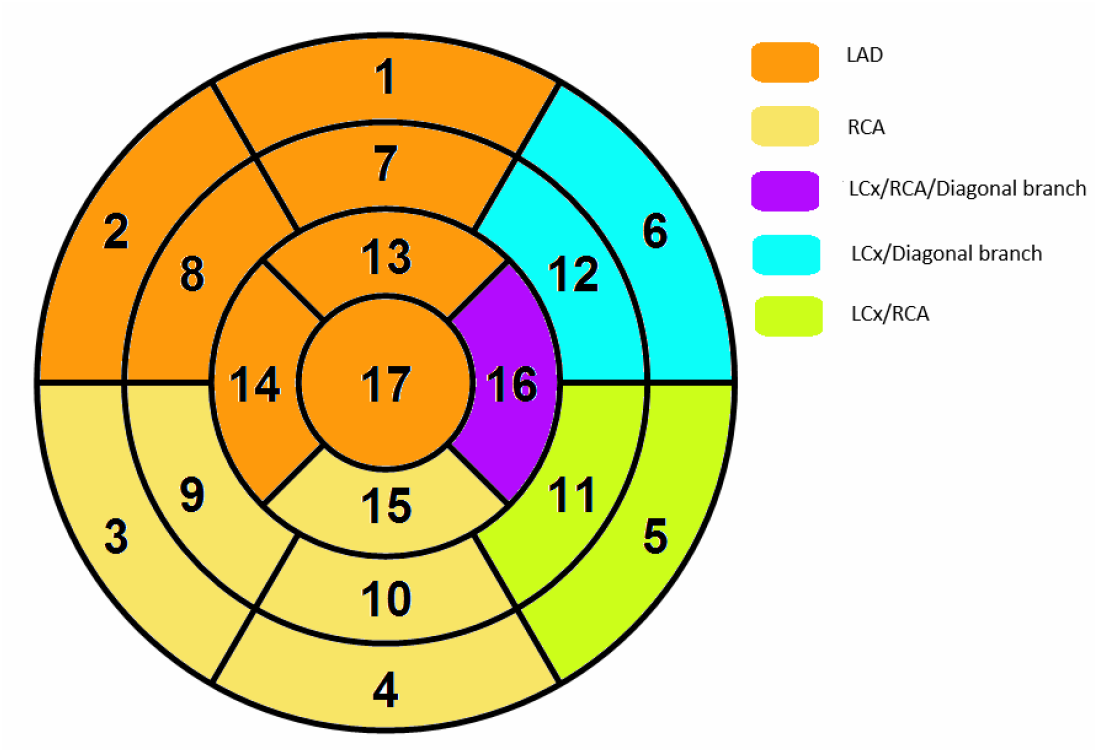
17 Segment AHA model

A total of 50 consecutive STEMI patients were analyzed. 78% were male, mean age was 63.9 ± 11.7, mean body mass index was 31.9 ± 6.0, mean global longitudinal strain was -11.3% ± 3.0% with ejection fraction of 46.4% ± 11.1%. LV AutoStrain analysis correctly identified the culprit vessel in 48 of 50 cases (96%), whereas routine clinical interpretation correctly identified the culprit vessel in 35 of 50 cases (70%). The only two cases not correctly identified by strain were confounded by a prior LAD infarct (RCA culprit) and left bundle branch block. This difference in diagnostic accuracy was statistically significant, with a p-value of 0.001. LV AutoStrain demonstrated a 100% accuracy rate in predicting LAD and LCx lesions, outperforming clinical interpretation, which achieved accuracy rates of 71% and 25%, respectively. The corresponding p-values were 0.009 and 0.143. LV Autostrain demonstrated a 91% accuracy rate in predicting RCA lesions, compared to 77% with clinical interpretation with a corresponding p-value of 0.412. Lastly, LV AutoStrain successfully detected LAD and LCx lesions with 100% accuracy, regardless of whether the echocardiographic image quality was most or least limited. These findings are highlighted in the graph illustrated in figure 3.

**Fig 3.**
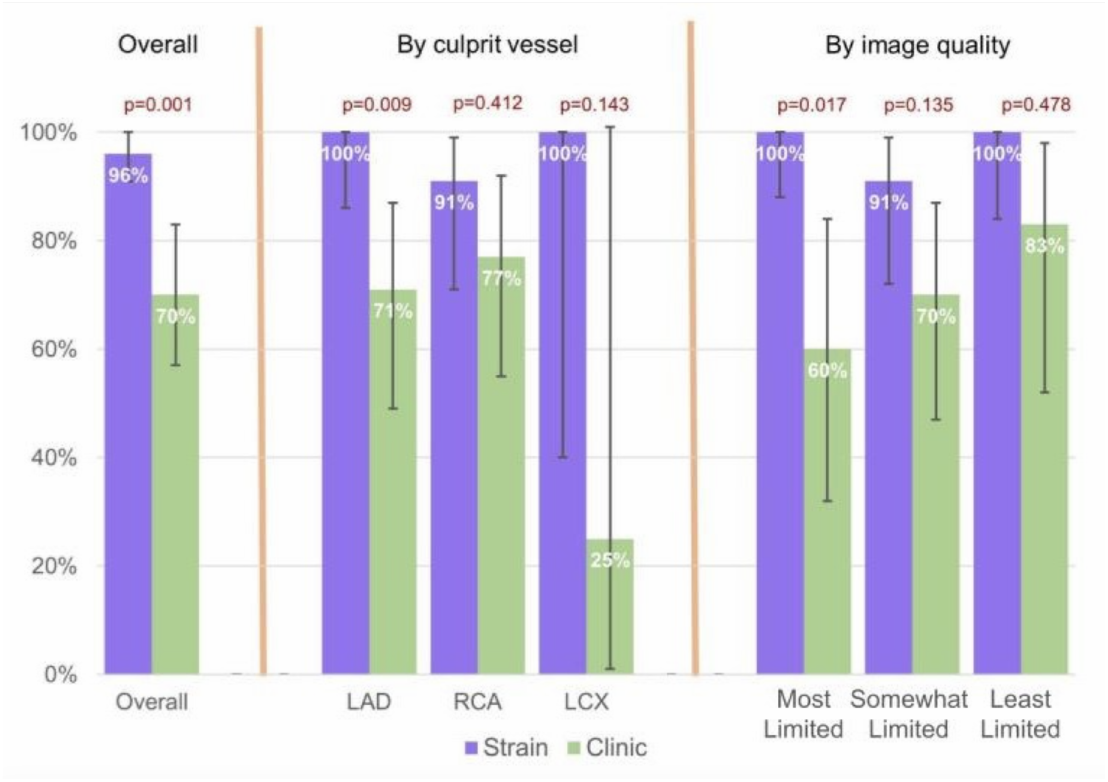
Comparing accuracy of longitudinal strain and routine clinical interpretation of echocardiograms according to culprit vessel and image quality

This study suggests that even low-quality echo images still contain valuable data that can be measured with longitudinal strain in order to identify regional deformation abnormalities. Indeed, strain, using very suboptimal imaging data, better predicted the culprit vessel than the standard clinical care interpretations. All echo studies with the basal insertion points and apical cap were included. The adjacent basal and apical segments were often suboptimally visualized. These criteria were broad and feasible in over 90% of screened studies. Our findings show strain to be a robust tool for identifying regional wall motion abnormalities, strongly supporting the use of strain in all patients. Strain performance did not change according to the vessel analyzed or the image quality. Routine clinical interpretation did show a non-statistically significant (p=0.139) trend toward improved accuracy as image quality improved though even in the “somewhat limited” studies, strain accuracy was numerically higher.

The TomTec AutoStrain tool is specifically designed to delineate the subendocardium. Limited subendocardial visualization is common with low-quality images. In this study, it is likely that suboptimal imaging led to the inclusion of the subepicardium in the region of interest. Subendocardial and subepicardial longitudinal strains are often closely correlated due to the transmural extent of many infarcts. Similar correlations between subendocardial and subepicardial strain have been observed in other cardiac pathologies, including hypertrophic cardiomyopathy and myocarditis [5,6]. Strain could potentially yield valuable measurements even in suboptimal imaging for other cardiac conditions with transmural involvement, thereby expanding its clinical utility.

The high accuracy of strain measurements will be particularly valuable in cases of myocardial infarction with nonobstructive coronary arteries (MINOCA). By using strain to identify regional wall motion abnormalities in a coronary vascular distribution, providers can more accurately identify probable MINOCA cases, refer for confirmatory MRI imaging and appropriately treat patients. As was observed in this work with 30% of the standard clinical care echo interpretations, regional wall motion abnormalities are often not identified. This increases the chances of MINOCA cases being misdiagnosed as nonischemic myocardial injury.

The 4AS parameter was highly accurate in predicting STEMI culprit vessels. This work adds to the prior research utilizing 4AS in patients with chest pain which had identified a cutoff value of >-15% for identifying pathology. All 4AS values in our work met this cutoff criteria, providing external validation of this cutoff.

Routine clinical echocardiogram interpretations were not standardized, making them susceptible to variability based on the reader’s level of expertise. Despite this limitation, GLS has previously demonstrated greater reproducibility in evaluating LV function compared to LVEF, regardless of the interpreter’s echocardiographic training [7]. This reinforces the reliability of strain analysis, even in the context of operator variability or suboptimal imaging quality. Furthermore, the inclusion of interpretations from multiple physicians provides a broad representation of diverse clinical perspectives, reflecting real-world practice. Additionally, in one case, LBBB led to the misidentification of the culprit vessel based on 4AS criteria. LBBB induces asynchronous myocardial contraction, which can confound strain-guided regional wall motion assessment.

Longitudinal strain better predicted the culprit vessel in STEMI patients with poor-quality echocardiograms compared to routine clinical interpretation. Despite traditionally held concerns that strain analysis on poor-quality images may be unreliable, this study suggests that strain can effectively utilize poor quality images to provide clinically valuable data. Given its significant improvement in diagnostic accuracy over standard care, integrating strain measurements into routine imaging workflows is highly recommended.

## Data Availability

All data produced in the present study are available upon reasonable request to the authors

## Acknowledgements

No relevant acknowledgements

